# Modelling the epidemic trend of the 2019-nCOV outbreak in Hubei Province, China

**DOI:** 10.1101/2020.01.30.20019828

**Authors:** Lizhe Ai

## Abstract

As of 8am 30^th^ January (Beijing Time) 2020, Approximate 8000 cases across the world have been confirmed. It’s necessary to simulate epidemic trend of the 2019-nCOV outbreak in Hubei Province, the hardest-hit area. By SEIR simulation, the predicted epidemic peak in Hubei will be within 28^th^ January 2020 to 7^th^ February 2020, up to 7000-9000 infectious cases in total. The estimate above was based on some assumptions and limitations exited.

## Background and current epidemic situation

In December, 2019, a series of pneumonia cases infected by a novel 2019-nCOV coronavirus emerged in Wuhan, Hubei Province, China. 2019-nCOV has demonstrated comparable transmissibility with infamous SARS and MERS, all belonging to the family Coronaviridae [1,2]. Surging with worrisome speed. As of 8am 30^th^ January (Beijing Time) 2020, Approximate 8000 cases across the world have been confirmed, of which 2261 cases confirmed in Wuhan City and 2325 cases confirmed in other cities in Hubei Province [3]. There was a mean 3-7day incubation period. And the most common symptoms of illness were fever, cough, and myalgia or fatigue [4]. Articles on modelling epidemic trend of 2019-nCOV have been published [5,6]. The predicted epidemic peak will be in late February 2020. Effective interventions implemented by Chinese Government, the epidemic will gradually die off in late March 2020.

## Transmission model

Due to Spring Festival, nearly 5 million people left off Wuhan before the traffic blockade January 10^th^ −22^th^, 2020. Fortunately, most of travelers above moved around in Hubei Province [7]. As the intense intervention implemented, the outbreak in provinces other than Hubei has been effectively under control (Figure1).

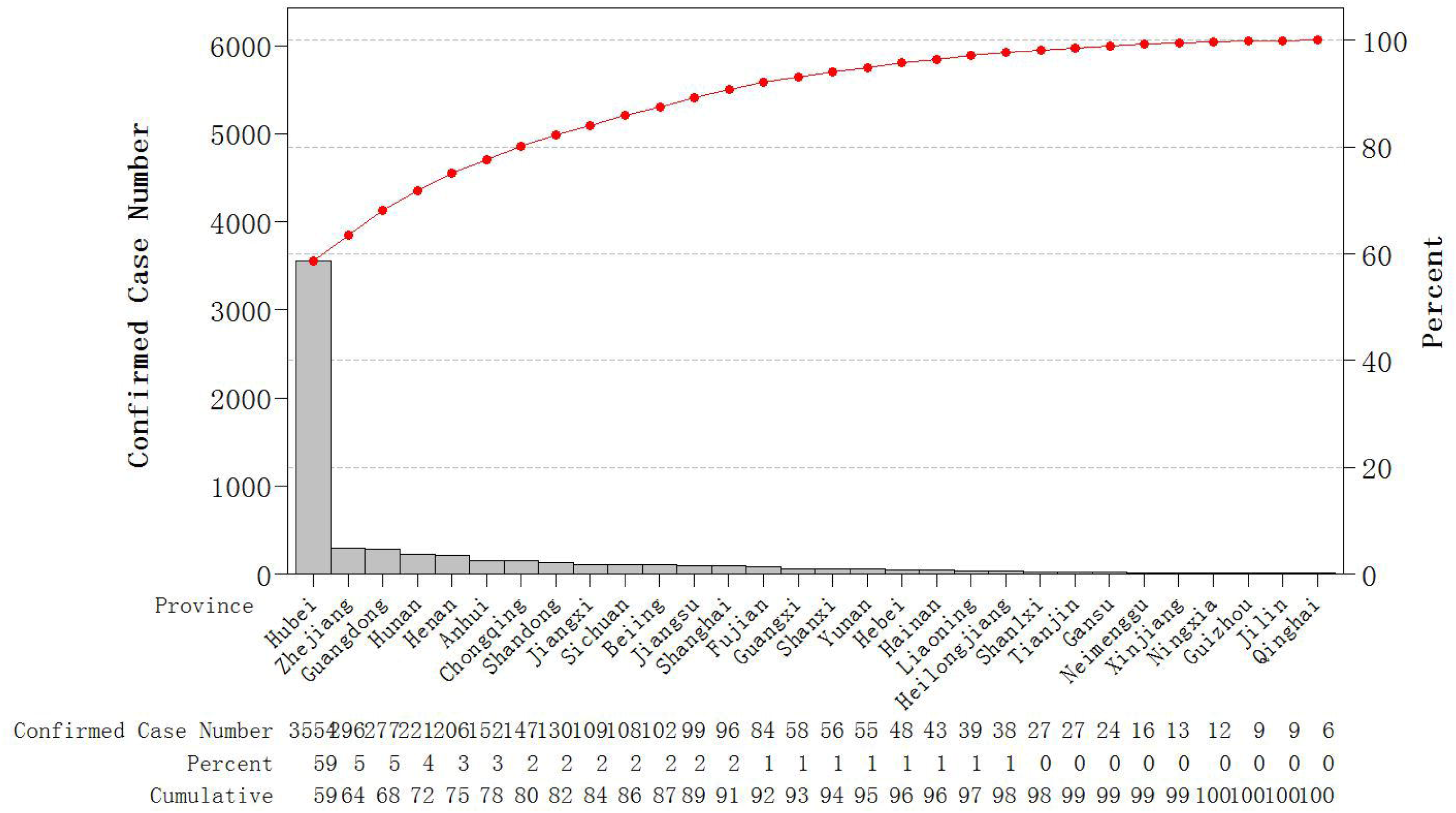

The epidemic trend focused on Hubei Province was simulated by a SEIR model. We estimated a total of approximate 5000 cases of 2019-nCOV in Hubei Province (mainly distributed in Wuhan City) by 22th January 2020. The transmission rate *β* in Hubei was calculated as 0.8 from 12^th^ December 2019 (the first 2019-nCOV case tracked) until 22^th^ January 2020 (traffic blockade started). Under the intense intervention, we assumed the transmission rate decreased to 0.005-0.1. With the assumption of no resurges resulted by virus mutation or unknown source of infection, the epidemic peak in Hubei will be within 28^th^ January 2020 to 7^th^ February 2020, up to 7000-9000 infectious cases in total.

## Discussion

Compared with SARS in2003, the emergency response capabilities of Chinese government have been improved. Interventions or relevant policies, such as free treatment for 2019-nCOV infectious patients, wildlife trade ban during the plague, traffic control, have been implemented rapidly. All models are wrong, but some are useful! The estimate above was based on some assumptions and limitations exited. Affected by Spring Festival travel, the epidemic has spread rapidly and trend is difficult to simulate. And further investigations on potential spatiotemporal transmission pattern are warranted.

## Model and Method

### PartI

The total number of cases in Hubei Province by 22th January 2020 is given by [5]:

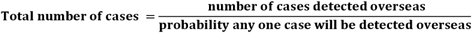

Where the probability any one case will be detected overseas (*p*) is given by:

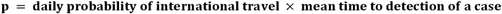

The daily probability of travel is calculated by:

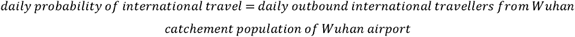

The mean time to detection can be approximated by:

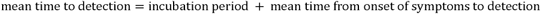

We set parameters and initial values as follows

Daily international passengers travelling out of Wuhan International Airport = 3301

Effective catchment population of Wuhan International Airport=19 million Detection window (days) =10 days

The Exported number of confirmed cases by 22th January 2020 = 9

Ignoring sporadic cases in cities other than Wuhan, we estimated 5000 cases of 2019-nCOV in Hubei Province by 22th January 2020.

### PartII

We proposed a SEIR model to simulate the transmission and trend of 2019-nCov in Hubei Province [6]. The population was divided into four compartments: susceptible individuals (S), asymptomatic individuals during the incubation period (E), confirmed infectious individuals (I), and recovered individuals (R). *β* denoted the mean person-to-person transmission rate per day in the absence of control interventions. Individuals in the incubation period progressed to the infectious compartment at a rate k. Infectious individuals recovered and died at the rate *γ*

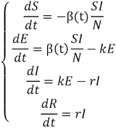

### Simulation 1

We set parameters and initial values as follows:

Average incubation time (k) =7;

Average illness duration (*r*) =10;

Population (N) =19million;

Susceptible individuals (S) =19million minus 1;

Symptomatic individuals during the incubation period (E) =0;

Confirmed infectious individuals (I) =1;

Recovered individuals (R) =0;

By simulation, when *β* approximate 0.8, the total number of case would be up to 5000 by 22th January 2020. Meanwhile, there were 10000 exposed people (E) and 1800 recovery people (R).

### Simulation 2

We set parameters and initial values as follows

Average incubation time (k) =7

Average illness duration (*r*) =10

Population (N) =59.17million;

Transmission rate (*β*) =0.005 to 0.1 by 0.005

Susceptible individuals (S) =59.17 million minus (E+I+R)

Symptomatic individuals during the incubation period (E) =10000

Confirmed infectious individuals (I) =5000

Recovered individuals (R) =1800

By simulation, the epidemic peak in Hubei will be within 28th January 2020 to 7th February 2020, up to 7000-9000 infectious cases in total (Figure 2).

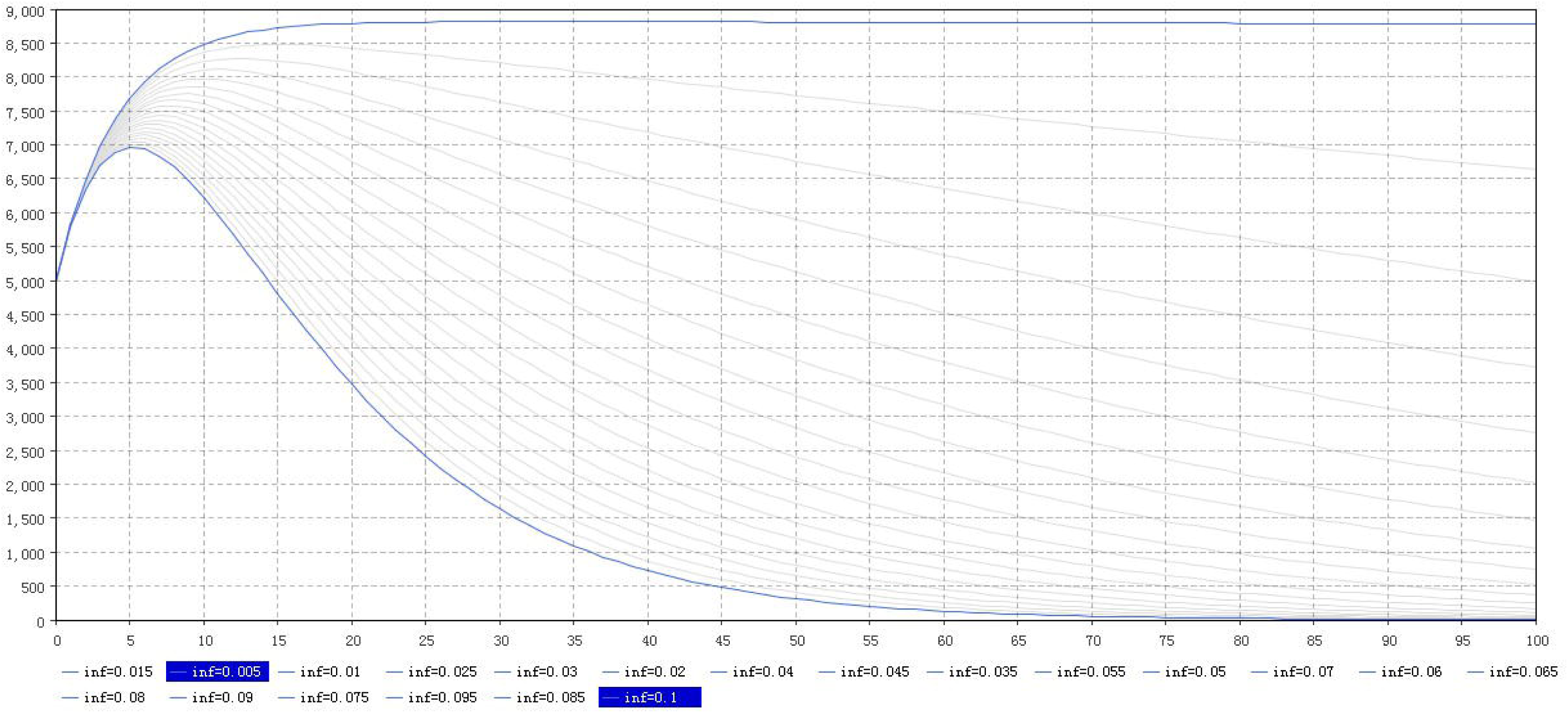

## Data Availability

all the data in the manuscript were collected from National Health Commission of the People’s Republic of China and published papers (including preprint)

## References

1. World Health Organization. Novel coronavirus (2019-nCoV). Situation report 3. 237 2020.

2. Wu P, Hao X, Lau EHY, Wong JY, Leung, K S M, Wu JT, et al. Real-time tentative assessment of the epidemiological characteristics of novel coronavirus infections in Wuhan, China, as at 22 January 2020. Eurosurveillance. 2020; 25: 2000044.

3. National Health Commission of the People’s Republic of China. Outbreak notification. http://www.nhc.gov.cn/xcs/yqtb/202001/5d19a4f6d3154b9fae328918ed2e3c8a.shtml

4. World Health Organization. Clinical management of severe acute respiratory infection when novel coronavirus (nCOV) infection is suspected: Interim guidance

5. Imai N, Dorigatti I, Cori A, Riley S, Ferguson NM. Report 1: Estimating the potential total number of novel Coronavirus (2019-nCoV) cases in Wuhan City, China [Internet]. 2020 [cited 23 Jan 2020]. Available: https://www.imperial.ac.uk/mrc-global-infectious-disease-analysis/news--wuhan242coronavirus

6. Mingwang Shen, Zhihang Peng, Yanni Xiao, Lei Zhang. Modelling the epidemic trend of the 2019 novel coronavirus outbreak in China. bioRxiv 2020.01.23.916726

7. Bjnews. Baidu map data shows that Wuhan’s population recently moved out of cities mostly in Hubei Province. http://www.bjnews.com.cn/finance/2020/01/23/678407.html

